# Did people really drink bleach to prevent COVID-19? A tale of problematic respondents and a guide for measuring rare events in survey data

**DOI:** 10.1101/2020.12.11.20246694

**Authors:** Leib Litman, Zohn Rosen, Cheskie Rosenzweig, Sarah L. Weinberger-Litman, Aaron J. Moss, Jonathan Robinson

**Author notes:** Correspondence concerning this article should be addressed to Leib Litman.

## Abstract

Society is becoming increasingly dependent on survey research. However, surveys can be impacted by participants who are non-attentive, respond randomly to survey questions, and misrepresent who they are and their true attitudes. The impact that such respondents can have on public health research has rarely been systematically examined. In this study we examine whether Americans began to engage in dangerous cleaning practices to avoid Covid-19 infection. Prior findings reported by the CDC have suggested that people began to engage in highly dangerous cleaning practices during the Covid-19 pandemic, including ingesting household cleansers such as bleach. In a series of studies totaling close to 1400 respondents, we show that 80-90% of reports of household cleanser ingestion are made by problematic respondents. These respondents report impossible claims such as ‘recently having had a fatal heart attack’ and ‘eating concrete for its iron content’ at a similar rate to ingesting household cleaners. Additionally, respondents’ frequent misreading or misinterpreting the intent of questions accounted for the rest of such claims. Once inattentive, mischievous, and careless respondents are taken out of the analytic sample we find no evidence that people ingest cleansers to prevent Covid-19 infection. The relationship between dangerous cleaning practices and health outcomes also becomes non-significant once problematic respondents are taken out of the analytic sample. These results show that reported ingestion of household cleaners and other similar dangerous practices are an artifact of problematic respondent bias. The implications of these findings for public health and medical survey research, as well as best practices for avoiding problematic respondents in surveys are discussed.

## Introduction

Surveys are one of the most common sources of data in social science (Singleton & Straits, 2009; Given, 2008; Connelly, Gayle, and Lambert, 2016), political science (Groves et al., 2011; Wolf et al., 2016, Mutz and Kim, 2020), public health (Ezzati-Rice and Curtin, 2001; Belisario, et al., 2015) and medical research (Saczynski, McManus, and Goldberg, 2013; Safdar, Abbo, Knobloch, and Seo, 2017) - informing public policy, medical practice and public opinion. Despite the widespread use of survey research, self-report data has come under increasing scrutiny over the last ten years due to data quality concerns.

One of the major threats to validity in survey research comes from participants who are inattentive, respond randomly to survey questions (Kim et al, 2018; King, Kim, and McCabe, 2018; see Chandler, Paolacci and Hauser, 2020 for review), or are ‘mischievous’, providing responses that are intentionally false or misleading (Fan et al, 2006; Robinson-Cimpian, 2014; Kramer, Rubin and Coster, 2014; Fish and Russell, 2018; Kaltiala-Heino and Lindberg, 2019; Li, Follingstad. Campe, and Chahal, 2020; see Cimpian et al., 2020 for a review). Inattentive and mischievous respondents (collectively referred to in this report as ‘problematic respondents’) can bias the results of surveys by dramatically inflating point estimates and creating illusory associations. In the current study we examine how problematic respondents can bias estimates of health-related behaviors. We additionally examine how rare events in public health research are particularly vulnerable to problematic respondent bias. In particular, we examine findings of recent survey data which indicate that Americans began to engage in highly dangerous cleaning practices in response to the COVID-19 pandemic (Gharpure et al., 2020). Such reported practices included drinking bleach and household cleaner to prevent COVID-19 infection.

Estimates of rare event frequencies, such as the ingestion of household cleansers, are particularly prone to problematic-responder bias (Robinson-Cimpian, 2014; Kennedy et al., 2020). The goal of the present study is to examine whether the rate of reported dangerous cleaning practices and the relationship between dangerous cleaning practices and health outcomes was overinflated in the Gharpure et al (2020) study due to inattentive and mischievous respondents. Another goal of this paper is to examine several different approaches to reducing problematic respondent bias and then demonstrate how, through their proper application, data quality can be vastly improved when collecting data from samples of survey respondents online.

## Background

Evidence that mischievous respondents can alter the outcomes of surveys began to accumulate as early as the 1970s. Having observed unusual patterns in survey data on self-reported illicit drug use, Petzel, Johnson, and McKillip (1973) began to suspect that some respondents were exaggerating their drug use in their self-reports. To examine this issue, they created a paradigm for catching potentially problematic respondents which involved incorporating questions about a fictitious drug in the survey. They found that 4% of people reported using a non-existent drug. They also observed that people who reported using the fictitious drug differed from the remaining sample on most other questions in the survey. Those who reported using a fictitious drug were much more likely to report using other drugs and to diverge from other respondents on virtually all other questions about drug use. The latter finding strongly suggested a general propensity toward acquiescence bias among these respondents.

A similar approach to identifying problematic respondents was used in a nationwide school-based study in Norway, in which close to 12,000 participants responded to questions about drug use. Pape and Strovoli (2006) found that respondents who reported buying and using the fictitious drug “Zetacyclin” also reported to be disproportionately heavy users of other drugs such as heroin and LSD. Because heroin and LSD use is a relatively rare event, excluding these respondents from the analytic sample made a critical difference for inferences about the nation-wide incidence of drug use.

Some of the most compelling demonstrations of the dramatic impact that problematic respondents can have on surveys come from the national Longitudinal Study of Adolescent Health (Add Health). Add Health uses multiple measurement methods, including surveys, in-person interviews and interviews with parents, which allows for survey responses to be directly cross-referenced with in-person interviews. Fan et al., (2006) reviewed multiple instances where in-person interviews directly contradicted survey responses. For example, 20% (176 out of 863) of respondents falsely reported not being born in the US, which was detected during the in-person interview. Nineteen percent (88 out of 458) of adolescents reported being adopted on a survey, but on in-person interviews their parents contradicted these claims indicating that their children were not adopted but were in fact their biological children.

In what is perhaps the most striking example of the potential of problematic respondents to completely invalidate the results of medical surveys, Fan et al., (2006) reported that out of 253 people who indicated on a survey that they had used an artificial limb for more than a year, 248 (99%) changed their response on a follow-up, in-person interview. Only 2 out of 253 people corroborated their prior claim made in the survey of having used an artificial limb when asked about it in person.

One clear pattern that emerges from studies that have examined problematic respondents is that once a respondent provides a false response, they are more likely to provide other false and problematic data. In other words, demonstrably false responses are typically a good indication that the entire survey should be treated with suspicion and that such participants should be considered for exclusion from the analytic sample. For example, people who falsely reported being adoptees or having a false limb were also less likely to be consistent when providing demographic information including gender, age, and race. While the correlation between the respondents’ age provided on the survey and their age determined from an at home interview was above .95 for people who did not provide false data, that correlation was .41-.47 for people who provided false reports. Further, the participants who provided false information also endorsed extreme responses on a wide range of behaviors such as alcohol consumption, leading to inflated and illusory between-groups disparities. A reanalysis of the Add Health data after having taken these respondents out of the analytic sample led the authors to conclude that several original reported disparities were “substantially overstated,” and led to retractions of published reports (see Cimpian and Timmer, 2020; Fan et al, 2006).

In contrast to the Add Health dataset, in which survey responses can be confirmed with in-person interviews, the responses of most surveys cannot be directly verified by in-person observation. To combat problematic respondent bias in surveys where responses cannot be directly verified, numerous data validity screening techniques have been developed which either identify mischievous and inattentive respondents prior to the survey and prevent them from participating in a survey (Chandler et al., 2019), or identify such respondents within a survey and remove their data from the analytic sample (Cornell, et al., 2012; Ward and Pond III., 2015; Litman, Robinson and Rosenzweig, 2015; Shukla and Konold, 2018; Kim et al., 2018; Steedle, Hong and Cheng, 2019; Leiner, 2019).

Using these techniques, it has become increasingly common to reanalyze extant survey data while controlling for problematic respondent bias in order to better understand how survey results in specific research areas may have been influenced by such bias (see Cimpian and Timmer, 2019). These efforts have helped to reveal that problematic respondents can drastically attenuate results, at times leading researchers to conclude that previously established findings lack validity (Cimpian and Timmer, 2019; Fan et al., 2002; Fan et al., 2006). While reexamination of previously reported survey results in light of problematic respondent bias can be viewed within the larger effort to improve replication reliability in the social sciences, even greater emphasis should be placed on examining results that have direct implications for public health and public policy (Vriesema and Gehlbach, 2019).

Problematic respondents are not bound to specific modalities of survey sampling, such as specific national databases or particular online survey platforms, and are not limited to specific demographic populations. Rather, problematic respondent bias is an ubiquitous problem that requires mitigation in any type of survey (e.g.Cimpian et al., 2018; Fish and Russell, 2018; see Chandler et al, 2019), leading to an almost unanimous call by researchers who study survey data quality for including rigorous methodology to support the validity of estimates drawn from survey data (Jia, Konold, Cornell and Huang, 2018).

One increasingly popular modality of collecting survey responses is via online opt-in panels. Such panels constitute more that 80% of currently conducted public opinion polls (Kennedy et al, 2020), and are increasingly being used for data collection in public health, political science, and social and behavioral sciences (see Chandler et al, 2019; Litman and Robinson, 2020 for a review). A large literature on opt-in panels indicates that a substantial percentage of problematic respondents exist on such panels (Downes-Le Guin, Mechling, & Baker, 2006 Courtright & Miller, 2011; Chakraborty, 2014; Hays, Liu, & Kapteyn, 2015; Chandler et al., 2019; Smith, Roster, Golden, & Albaum, 2016; Kees, Berry, Burton, & Sheehan, 2017; Kennedy et al., 2020; Litman, Robinson, & Rosenzweig, 2020b). Estimates of problematic respondent bias magnitude on online opt-in panel platforms vary between 4-7% (Kennedy et al, 2020) and 30% (Chandler et al, 2019), although in some studies the magnitude of inattention has been as high as 50% (see Kees et al, 2017).

Critically, the problematic survey responses obtained on online panels are not random. The largest and most comprehensive study done to date to systematically examine problematic responses on online opt-in panels shows that responses tend to be systematically skewed toward positive answer choices (Kennedy et al, 2020). Specifically, this means that when provided with a yes/no response option, problematic respondents will be more likely to choose a ‘yes’ response over a ‘no’ response. A systematic yea-saying tendency, also known as acquiescence bias, creates a particularly grave concern for studies which aim to measure rare events. This is because even a small percentage of respondents who falsely answer yes to questions about rare events will make a non-existent phenomenon appear to be real. For example, it has been noted that even a small percent of problematic yea-saying can bias a political poll (see Kennedy et al., 2020). However, little research has directly addressed how such yea-saying bias stemming from problematic respondents in online panels can skew the results of public health studies attempting to identify rare behaviors and rare populations. The present study directly addresses this issue; specifically, we examine how incorrect ‘yes’ responses can artificially inflate estimates of rare public health-relevant behaviors in online opt-in samples.

### Present study

The Covid-19 pandemic has had a profound influence on daily health-related practices in the United States and around the world. The World Health Organization and the Center for Disease Control (CDC) have issued multiple behavior guidelines to help curb the spread of infection, including wearing a face mask and social distancing. Some of the most important health-related guidelines relate to cleanliness practices, including the need to wash hands thoroughly and often, and to avoid hand-to-face contact (CDC, 2020; WHO, 2020).

Previous research has shown that even before COVID-19, cleanliness and contamination concerns have led people to engage in a variety of cleaning practices to reduce the likelihood of infection, particularly surrounding food cleanliness (see Litman et al., 2018; Litman et al., 2019). At times, contamination concerns can lead people to engage in dangerous cleaning practices, such as overusing antimicrobial products that can lead to skin damage and cause other health problems (Larson, 2001). It is thus reasonable to expect that during the time of COVID-19, when fear of contamination and infection is very high, people will be even more likely to engage in a variety of cleanliness practices as they seek to protect their health. Indeed, fear and concern about COVID-19 infection may be so high during a pandemic that people may engage in cleanliness practices that are detrimental to health, or in extreme cases, may even be dangerous.

In a recent study, Gharpure et al., (2020) presented evidence that such dangerous cleaning practices appear to be happening at an alarming rate. In their online survey, Americans were asked if they had engaged in highly dangerous cleaning practices such as drinking or gargling disinfectants, household cleaning products, or soap to prevent Covid-19 infection. Their data revealed that during April of 2020, 39% of Americans engaged in at least one cleaning practice not recommended by the CDC. Such practices included 19% of people who applied bleach to food items; 18% who used household cleaning products on their skin; 10% who misted their body with a disinfectant; 6% who inhaled vapor from a cleaning product or disinfectant; and particularly alarming, a non-trivial percentage of people who ingested cleaning products, including 4% who reported drinking or gargling diluted bleach, 4% who reported drinking or gargling soapy water, and 4% who reported drinking or gargling a household disinfectant. Translating these percentages into population figures, these results imply that approximately ten million American adults were ingesting bleach, and twenty million American adults were ingesting at least one cleaning product.

The finding that Americans were engaging in dangerous cleaning practices at such high rates are alarming. It suggests that fear of COVID-19 contamination, coupled with a lack of knowledge about the dangers of such practices, are leading tens of millions of people to engage in behaviors that can damage their overall health. Indeed, the scale at which people are engaging in such practices may be revealing an area of public health concern that requires large-scale intervention (Gharpure et al., 2020).

However, there are several reasons that these results should be interpreted with caution. As in any study aiming to detect rare events using survey research, a combination of acquiescence bias, inattention, and mischievous responding - all factors that have been extensively documented to bias surveys both in and outside of online opt-in panels - can severely bias the estimates of such behaviors. For this reason we sought to examine whether reports of dangerous cleaning practices, and claims of ingestion of cleaning products in particular, can be attributed in part or in whole to problematic respondent bias.

Aside from the methodological concerns about bias in survey research, there are other important public health considerations motivating us to verify whether the ingestion of cleaning products is occurring at this alarmingly high rate. The medical literature does indeed document that people ingest cleaning products, and that at times such practices can lead to severe health complications including death (Williams et al, 2012). However, the occurrence of such practices is very rare, and when it does occur people are almost exclusively motivated to ingest cleaning products for their alcohol content rather than their perceived health benefits (Rayar and Ratnapalan, 2013). Presenting the occurrence of dangerous practices as being more widespread than they actually are is also of concern to public health. Decades of social norm research suggests that people’s behavior may be influenced by what is perceived to be normative (see Legros and Cislaghi, 2020). Experimental evidence suggests that social norms can alter risk-perception thereby influencing behavioral choices (Veflen et al, 2020), and increase vulnerability to misinformation (Myrick and Erlichman 2020). It is therefore possible that the few people who do engage in ingesting cleaning products for their alcohol content may be reinforced by the idea that millions of other people are also engaging in such practices. Therefore, if the ingestion of household cleansers is indeed being overreported, this may have the unintended consequence of reducing barriers associated with such practices.

Of course, it is also possible that people really did begin to ingest household cleaners due to increased concerns about safety during the time of Covid-19. If this is actually the case it is critical to verify that these practices really are occurring by disconfirming the hypothesis that such practices are being reported almost exclusively by problematic respondents. Thus, our overall goal in this study was to examine whether reports of dangerous cleaning practices such as ingesting household cleaners to prevent COVID-19 infection can be detected after controlling for problematic respondent bias. Specifically, we sought to systematically measure the magnitude of problematic respondent bias in influencing estimates of dangerous cleaning practices, with a focus on the ingestion of household cleaners including bleach, soap water, and household disinfectant. We aimed to determine what role problematic respondents may play on the reporting of these practices, and to more accurately measure how widespread such practices are.

### Detecting problematic respondents

In this study, several instruments were used to identify problematic respondents. All of these instruments, which are described in more detail below, have previously been shown to be effective at detecting different aspects of problematic responding and have been used to reduce various aspects of problematic respondent bias. Some of these instruments are optimized for detecting inattentiveness, others are optimized for detecting mischievous responding, while others are optimized for detecting acquiescence bias. In the current study we employ a combination of different instruments to address multiple known characteristics of problematic respondent bias.

#### Instrument 1 - Checks of attentiveness (Used in Study 1 and Study 2)

The first method we use was developed by Chandler et al., (2019), who specifically explored ways to reduce problematic respondent bias in online opt-in panels. Chandler et al, showed that up to 30% of data collected from online samples come from problematic respondents and that failing to eliminate such respondents from the analytic sample led to biased estimates of experimental effects and a dramatic misrepresentation of participants’ true behavior. Chandler et al (2019) used four screening questions aimed at checking for attentiveness and basic English language comprehension. Incorporating this screening methodology to detect problematic respondents and to exclude them from the analytic sample resulted in dramatic improvements in measurement accuracy.

#### Instrument 2 - Checks of mischievous responding (Used in Study 1 and Study 2)

The second method we used was originally developed by Petzel, Johnson, and McKillip (1973) and subsequently used by multiple other methodologists (e.g. Pape and Strovoli, 2006; Cimpian and Timmer, 2019; Chandler et al 2019). It involves incorporating questions within the survey about highly unlikely or impossible behaviors such as using a non-existent drug or ‘eating concrete for its iron content’ to identify problematic respondents.

#### Instrument 3 - Responsive verification as a way of reducing acquiescence bias (Used in Study 2)

Loftus et al (1990) showed that one way to increase accuracy in surveys responses is to ask respondents to report on behaviors of interest multiple times. Their study revealed that patients tend to overreport whether they have had a physical examination, as verified by patient records (also see Armstrong, Long and Shea, 2004). They also found that incorporating a second question to verify the original response significantly improves reporting accuracy. This occurs in part because it signals to participants the importance of the question to the researcher (see Loftus et al., 1990). In Study 2, we utilize this verification methodology, and additionally incorporate other verification questions that are based on the in-person interview techniques developed by Fan et al (2006), discussed above. While verifying responses via direct in-person interviews is impossible in online surveys, it is still possible to incorporate some level of verification by probing responses provided on a survey.

Responsive verification in our study involved getting the respondent to confirm their self-report of a rare event on a follow-up question embedded in a later part of the survey. This way, inattentive respondents have a second chance to correctly address the question, and respondents who may have mistakenly pressed the wrong button have a chance to correct their error. Additionally, we utilized open ended questions to understand more about the context and motivations behind the reported behavior. Overall, responsive verification can provide additional clarity and confidence to data collection and interpretation.

#### Instrument 4 - Validation of demographic information (Used in Study 2)

The final method we use to identify problematic respondents is based on Robinson-Cimpians’ (2014) method of validating demographic information. This method involves looking across reported demographics to find inconsistencies and exaggerated claims. Robinson-Cimpian developed a quantifiable metric for flagging problematic respondents based on an outlier analysis. Here, we utilize a similar approach by flagging demographic claims that are clearly implausible.

Study 1 used instruments 1 and 2 (checks of attentiveness and checks mischievous responding) to identify problematic respondents while Study 2 employed all four instruments.

### Study 1

#### Research objectives and hypotheses

Study 1 had several research objectives and hypotheses.

##### Hypothesis 1

The first objective of Study 1 was to examine the proportion of the seven dangerous cleaning practices (see Table 2) that come from problematic respondents. We hypothesized that the majority of reports of all seven dangerous cleaning practices come from problematic respondents. Further, we hypothesized that the lower the frequency of the reported behavior, the higher the proportion of affirmative responses will come from problematic respondents.

##### Hypothesis 2

The second objective was specific to the three of the most dangerous cleanser ingestion practices: 1) drinking or gargling bleach, 2) drinking or gargling disinfectant, and 3) drinking or gargling household cleaner. We hypothesized that the vast majority of individuals reporting ingesting cleansers would be identified by our instruments as being problematic. For these cleanser ingestion practices, we predicted that 80-90% of reports of ingesting household cleaner will come from problematic respondents.

##### Hypothesis 3

Our third objective was to demonstrate that respondents who are not identified as problematic by our instruments will report having ingested household cleansers at a very low rate. Specifically, our objective was to demonstrate a low false negative rate of the screening instruments. We expected that less than 1% of respondents who are not flagged by our instruments will report having ingested household cleansers.

##### Hypothesis 4

We additionally wanted to examine whether the presence of problematic respondents in surveys can make it appear that any behavior - no matter how implausible - will be reported with a base rate similar to the self-reported frequency of cleanser ingestion. We predicted that the confirmation frequency of implausible behaviors such as eating concrete, and “I have recently had a fatal heart attack”, will be similar to the confirmation frequency of cleanser ingestion.

##### Hypothesis 5

The fifth objective was to examine the correlation between dangerous cleaning practices and health outcomes (see Table 2 for the full list of health outcomes). We hypothesized that; a) there will be a high correlation between cleanliness practices and health outcomes, and b) taking problematic respondents out of the analytic sample will attenuate the association between cleanliness practices and health outcomes. As mentioned above, previous studies have shown that taking problematic respondents out of the analytic sample attenuates correlations (see Cimpian and Timmer, 2020). This is because problematic respondents tend to acquiesce to multiple unrelated survey items, artificially driving up correlations between unrelated events.. We thus hypothesized that the association between cleanliness practices and health outcomes will be very high among problematic respondents and will be very low among non-problematic respondents.

## Method

### Sample and survey design

A survey containing questions about cleanliness practices during the time of COVID-19 was fielded online to a National sample during the week of June 10th-June 17th. The sample of 600 respondents was matched to the U.S. Census on gender, age, race, and region (see Table 1 for the respondents’ demographics). The survey included questions about dangerous cleanliness practices people engaged in as a response to the COVID19 pandemic (e.g. Introductory question: “In the past month, which of the following cleaning behaviors have you or a household member engaged in to prevent coronavirus? Specific practice: Drank or gargled a household cleaner”. Response options: Yes/No), knowledge about safe cleanliness practices (e.g. Introductory question: “Which of the following have you heard is true about using household cleaning products (such as bleach or Lysol)? - Specific question: Household cleaning products should be kept out of the reach of children”. Response options: Yes/No), and questions about experiences with negative health effects due to the use of household cleaners (e.g. General question: “During the last month, have you experienced any of the following health effects due to using cleaners or disinfectants? Specific question: Breathing problems”. Response options: Yes/No). The complete survey including questions and response options is available in the Supplementary Materials.

**Table 1.** Basic demographics.

|  | Study 1(%) | Study 2 (%) |
| --- | --- | --- |
| Age | 50.0 (17.2) | 43.1 (17.8) |
| **Gender** |  |  |
| Female | 53 | 59 |
| **Race** |  |  |
| White | 71 | 67 |
| Black | 13 | 13 |
| American Indian or<br>Alaska Native | 2.2 | 2.2 |
| Asian | 8 | 8 |
| Native Hawaiian or<br>Pacific Islander | 0.2 | 0.8 |
| Multiracial/Other | 2.5 | 9 |
| **Hispanic** |  |  |
| Yes | 17 | 23 |
| **Highest Degree** |  |  |
| No college degree | 35 | 48 |
| Associate degree | 10 | 12 |
| Bachelor's degree | 31 | 23 |
| Graduate degree | 24 | 17 |
| **Region** |  |  |
| Northeast | 25 | 24 |
| Midwest | 21 | 23 |
| South | 35 | 31 |
| West | 19 | 17 |

The current study was a replication of the Gharpure et al. (2020) study, and we thus used the identical survey design, question wording, online sample provider, and generally attempted as much as possible to use the same sampling methodology as reported in the method section of the Gharpure et al. (2020) study.

### Screening methods

In Study 1, two screening instruments were used: 1) checks of attentiveness, and 2) checks of mischievous responding.

#### Attentiveness instrument

The attentiveness check instrument was based on the procedure developed by Chandler at al., (2019), which checks for attentiveness and basic English language comprehension. Chandler et al.’s, 2019 screening procedure consists of presenting four questions which have a target word and four response-option words. Participants are asked which response-option is most similar to the target. Chandler et al., showed that this instrument identified the vast majority of inattentive respondents in online samples, while having very low levels of false positives (i.e. almost all non-problematic respondents answer these questions correctly).

Here, we improved on the Chandler et al., methods by having refined and extensively tested each question in the instrument. The stimuli were generated by an associative semantic network algorithm, which assigned weights to word-pairs based on corpora of English language texts. Word-pairs were assigned weights based on the similarity and frequency with which they appear together. Of the four response options, the correct response was highly associated with the target (e.g apologize-mistake) and the other three had low associations with the target (e.g apologize-particle).

Only very common words that have a high frequency of occurrence in the English language were used as targets or response options so as to avoid education bias. Screening questions with a pass rate of 95% or above were used, as based on pilot testing conducted on independent participant samples. Using this approach a library of questions was developed so that different combinations of screening questions can be presented to different participants. This prevents bots and problematic respondents from learning correct responses to these questions, sharing them online, and creating scripts for response automation.

#### Mischievous responding instrument

The second instrument used in Study 1 was employed to detect mischievous responding. Participants were asked about events and abilities where only one of the responses could plausibly be the correct one (e.g. “Can you recall from memory the names of every single senator who ever served in the Senate’? (correct answer ‘No’), and ‘Did you ever use the internet’? (correct answer ‘Yes’). Each of these questions was likewise pre-tested to make sure that over 95% of attentive respondents answer in the expected way.

The instruments for detecting problematic respondents were presented at the end of the survey so as not to alter the number of problematic respondents entering the survey. Presenting the instruments at the beginning of the survey causes some problematic respondents to drop out prior to the survey (Chandler et al, 2019), and this would impact the accuracy of the measure of the effect that problematic respondents have on reports of dangerous cleaning practices.

### Analytic approach

Our first analytic goal was to compare the reported incidence of dangerous cleaning practices among respondents who were flagged by our instruments (problematic respondents) to those who were not flagged by our instruments (non-problematic respondents). Respondents were classified into a ‘problematic respondent’ group if they incorrectly responded to any of the items on instruments 1 and 2, and a ‘non-problematic respondent’ group if they answered all such items correctly. The reported incidence of dangerous cleaning practices was computed separately within those groups for all cleaning practices queried in the survey, allowing us to address hypotheses 1 - 4.

Hypothesis five aimed at comparing the association between engaging in dangerous cleaning practices and health outcomes between problematic and non-problematic respondents. For this analysis, a ‘dangerous practices’ score was computed by summing across all seven cleaning practices quarried in the survey. Likewise, a ‘health outcomes’ score was computed by summing across all six of the symptoms queried in the survey. The association between dangerous practices and health outcomes was explored with OLS regression, controlling for age, gender, race, education, and region of the country. Regression analyses were conducted separately for problematic and non-problematic respondents. We predicted that the percent of variance of health outcomes explained by dangerous cleaning practices, as measured by the R^2^, would be substantially higher among problematic respondents, and would be either very small or non-significant among non-problematic respondents.

## Results

Across the full sample, we observed that 3.8% of respondents reported drinking or gargling diluted bleach solution, 4% reported drinking or gargling soapy water, and 4.7% reported drinking or gargling household cleaner.

As predicted, the reported rate of engaging in these dangerous cleaning practices was similar to the reported rate of other survey items that are impossible. Specifically, 5.8% of respondents reported having “suffered a *fatal* heart attack” and 3% reported that they have never used the Internet. These results show that a confirmation bias base rate of between 3-6% can be expected for any question, no matter how implausible.

In the next analysis, we examined the differences in self-reported cleaning practices across all cleaning questions between those who passed the screener and those who did not pass the screener. Overall 76.7% (460/600) of respondents passed the screener, and 23.3% (140/600) did not pass the screener. As shown in Figure 1, the results were striking, showing that people who did not pass the screener reported engaging in dangerous cleaning behaviors much more frequently than people who passed the screener. For each of the cleaning practices examined in the survey, the likelihood of a ‘yes’ response was between 400% - 2400% higher among problematic respondents (see Figure 1).

**FIGURE 1.**
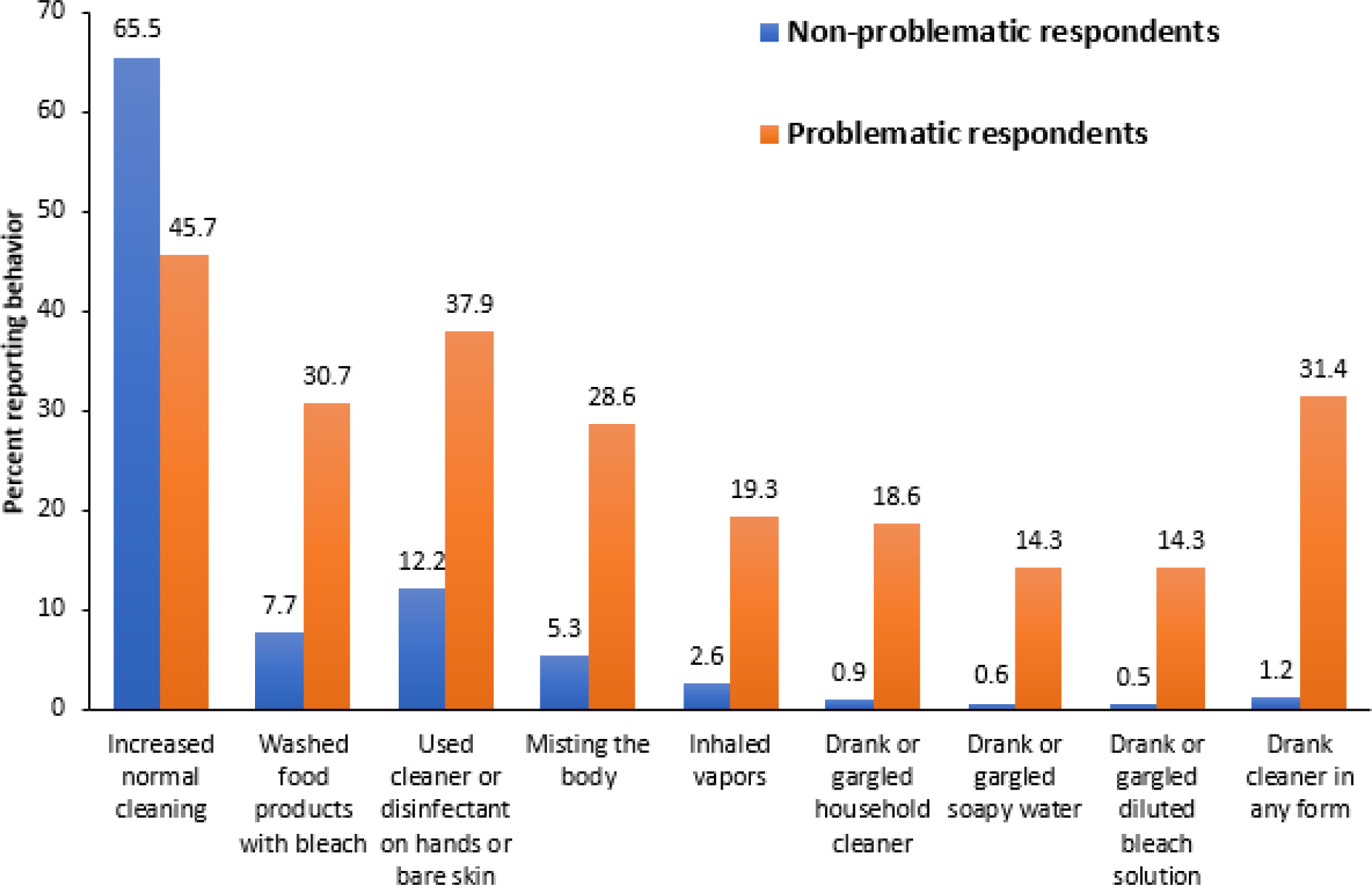
Comparisons of problematic and non-problematic respondents’ reports of cleaning and disinfection practices since April, 2020.

The highest discrepancy between problematic and non-problematic respondents was observed for household cleanser ingestion (i.e. drinking/gargling household cleaner, soapy water, or diluted bleach). Overall, 31.5% of problematic respondents reported engaging in at least one of these practices. In comparison, 1.2% of non-problematic respondents reported engaging in at least one of these cleaning practices.

Looked at another way, 88% of those who reported drinking or gargling household cleaner, soapy water, or diluted bleach, were identified as problematic respondents. In comparison, among those who did not report drinking or gargling household cleaner, soapy water, or diluted bleach, 17.5 % were identified as problematic respondents (see Figure 2).

**FIGURE 2.**
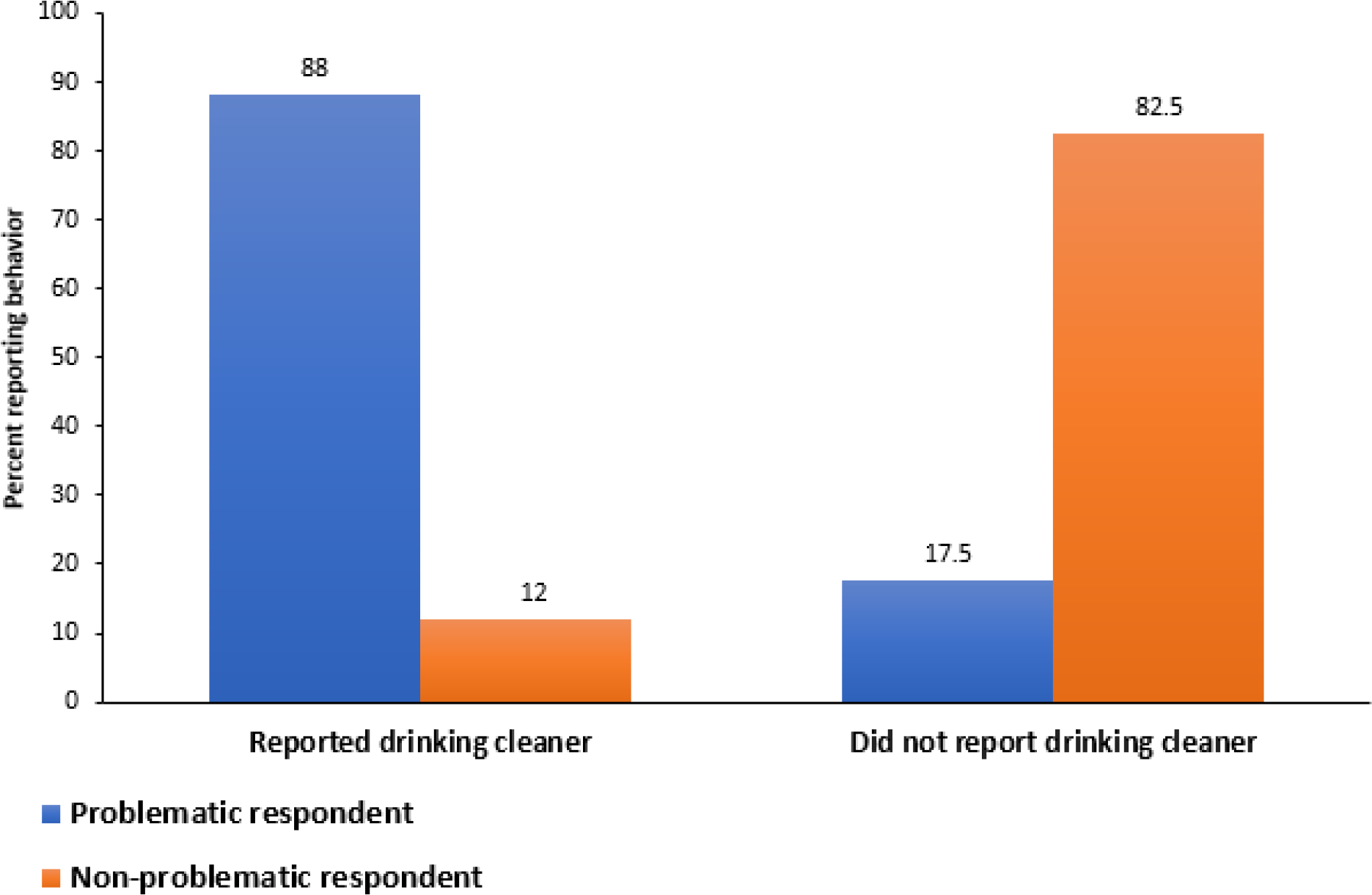
Probability of being labeled a problematic respondent based on whether participants reported drinking household cleanser.

Equally as important, the opposite pattern was observed for increases in typical non-dangerous cleaning practices. It is expected that normal cleaning behaviors such as hand washing would increase during the pandemic driven by public health recommendations. Problematic respondents under-reported increases in normal cleaning behavior by close to 20%. Thus, problematic respondents severely overreport high-risk cleaning practices and underreport regular cleaning practices.

In the next analysis we evaluated the association between dangerous cleaning practices and health outcomes using OLS regression-based modeling. We began by looking at a simple bivariate association between the number of dangerous cleaning practices reported on the survey and the number of reported health symptoms. We found that reported dangerous cleaning practices accounted for 18.1% of the variance in the number of reported health symptoms among the full sample, F (1, 598) = 132.3, p. < 001.

The association between the number of dangerous cleaning practices reported on the survey and the number of reported health symptoms may, however, be inflated by the tendency of problematic respondents to systematically select a ‘yes’ response across unrelated sets of questions. To address this potential confound, we modeled the association between the number of dangerous cleaning practices reported on the survey and the number of reported health symptoms separately for the identified problematic and non-problematic respondents.

We found that the association between reported dangerous cleaning practices and health outcomes was much lower among non-problematic respondents. Specifically, among problematic respondents 27.2% of the variance in the number of reported health symptoms was explained by reported dangerous cleaning practices, F (1, 138) = 132.3, p. < 001; in contrast, among non-problematic respondents, 3.5% of the variance in the number of reported health symptoms was explained by reported dangerous cleaning practices, F (1, 458) = 17.6, p. < 001.

## Discussion

Using instruments for detecting inattentiveness and mischievous responding to cluster people into problematic and non-problematic respondent groups, we observed that the vast majority of dangerous cleaning practices were reported by problematic respondents across all seven cleaning questions. Among all questions, the ones with the highest proportion of problematic respondents were practices involving the ingestion of household cleansers. Eighty eight percent of reports of these practices were made by problematic respondents. These data demonstrate that the rate of dangerous cleaning practices is largely an artifact of problematic respondent bias and that survey studies are vulnerable to such bias, particularly when attempting to detect rare events.

There was still, however, a small minority of respondents who were not identified as problematic who did report ingesting household cleaners. While ingestion of household cleaners was reported at a very low rate among non-problematic respondents (0.8%, 0.6% and 0.6% for drinking/gargling household cleaner, soapy water and diluted bleach respectively), a key question is whether the two instruments used in Study 1 were sensitive enough to detect all problematic respondents. More specifically, is it possible that a more sensitive test can reveal potential problems with responses that come from participants who passed the first set of screeners?

One possibility is that even respondents who pass screens of inattentiveness and mischievousness may at times lose focus, misunderstand the intent of specific questions, or mistakenly press the wrong button when answering a question. To verify that the type of respondents who were labeled as non-problematic in Study 1 really did ingest household cleansers, in Study 2 we employed two additional instruments: responsive verification, and validation of demographic information. Responsive verification consists of three steps: 1) Respondents who indicate that they ingested household cleaner were presented with a follow-up question asking them to verify their response, 2) Those who verified that they ingested household cleaner were asked whether they did so intentionally or unintentionally, and 3) Respondents were asked to describe what had occurred in an open-ended format. The purpose of the open-ended question was to understand more about the specific details and context for the reported behavior and the motivations behind it.

### Study 2

#### Research objectives and hypotheses

This study had several research objectives and hypotheses.

##### Hypothesis 1

Our first objective was to replicate the finding of Study 1, that most reports of cleanser ingestion come from problematic respondents. We hypothesized that, as in Study 1, 80-90% of reports of ingesting household cleaner will come from problematic respondents.

##### Hypothesis 2

Our second objective was to replicate the finding from Study 1 that respondents who are not red flagged by our instruments report having ingested household cleansers at a very low rate. We hypothesized that less than 1% of validated respondents who are not flagged by the attentiveness and mischievous responding instruments will report having ingested household cleansers.

##### Hypothesis 3

Our third objective was to examine whether reports of ingesting cleansers among respondents who passed attentiveness and mischievousness screens are due in part to misunderstanding the questions. We hypothesized that among those respondents who pass the attentiveness and mischievous responding screener and still report ingesting household cleaner, the majority will either fail to confirm that they did so on a follow up question, or indicate that they ingested household cleaner by accident, rather than to prevent Covid-19 infection.

##### Hypothesis 4

The fourth goal was to understand more about people’s practices and motivations by having them answer an open-ended question about their reported behavior. We hypothesized that we would see few if any cogent open-ended descriptions of ingesting household cleaner. In contrast, we expected to see informative open-ended responses of other practices, such as washing food such as fruits with bleach.

##### Hypothesis 5

As in the first study, we wanted to examine the correlation between health and dangerous cleaning practices (See Table 2). To do that we examined this correlation after having excluded respondents who failed to verify that they ingested household cleaner. We hypothesized that taking the problematic respondents and those who did not confirm having ingested household cleaner out of the analytic sample will attenuate this association to an even greater extent than the attenuation observed in Study 1.

**Table 2.**
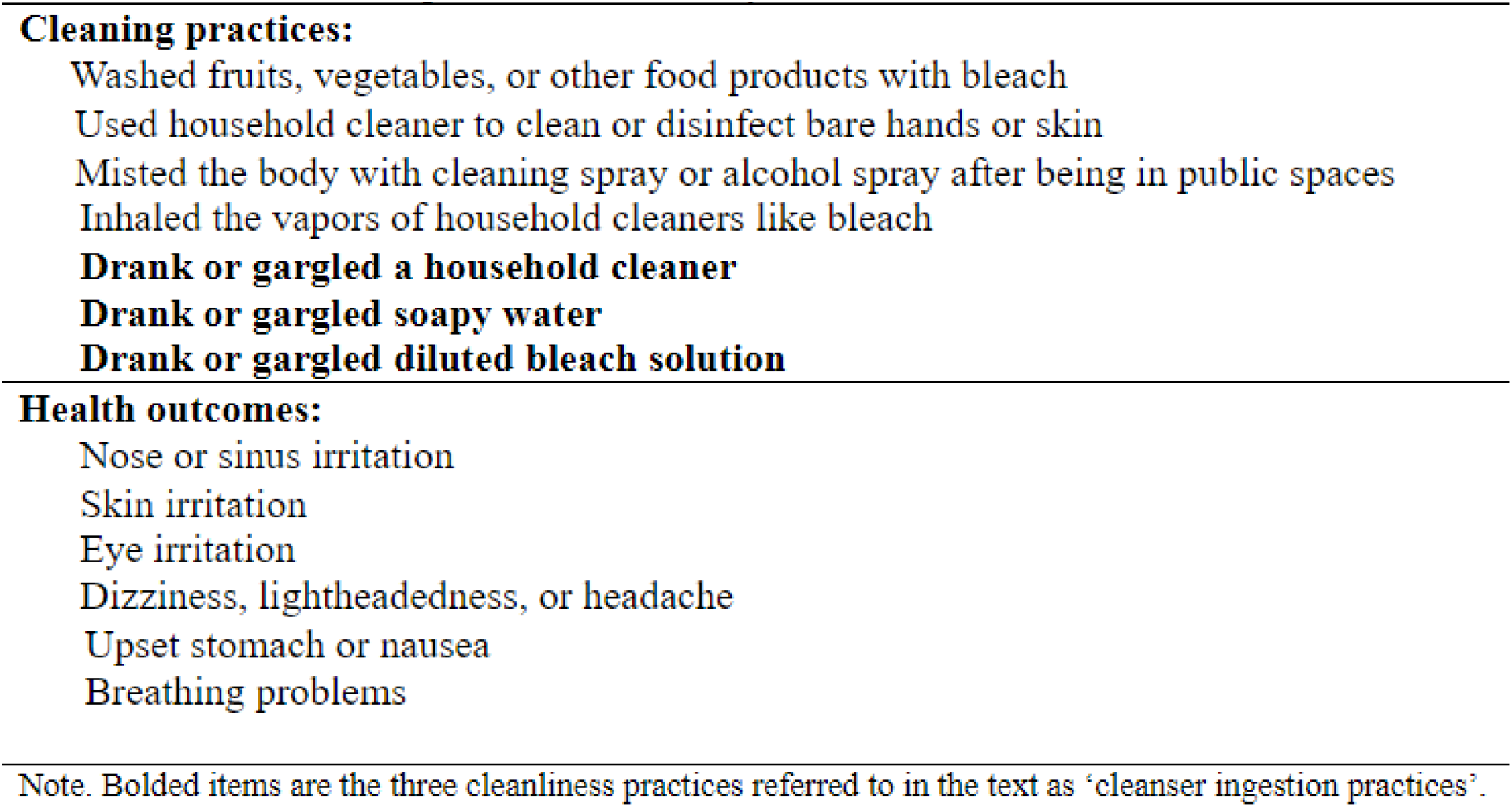
Practices and outcomes queried on the survey

## Method

### Sample and survey design

The overall survey content and participant sourcing methodology was identical to study 1, with two notable exceptions. First, the attentiveness and mischievous responding instruments were placed at the beginning of the survey rather than at the end. Second, because this study was conducted a few weeks after the first, the wording of the introductory questions was changed to “*Since the start of the Covid-19 pandemic in April*, which of the following cleaning behaviors have you or a household member engaged in to prevent coronavirus? Third, when respondents indicated engaging in dangerous cleaning practices, a series of follow up questions were asked to verify their response as part of the responsive verification protocol. Responsive verification prioritized the three questions about ingesting household cleansers. Specifically, follow-up questions were asked whenever respondents indicated that they ingested cleanser, bleach, or soapy water. For those respondents who did not report ingesting household cleaner but did report engaging in other dangerous cleaning practices, follow up verification questions were also asked about those practices as well.

The survey was fielded online to a National sample during the week of July 27th-July 31st. The sample of 688 respondents was matched to the U.S. Census on gender, age, race, and region (See Table 1).

### Screening methods

In addition to the instruments used in Study 1, we additionally used the responsive verification protocol and validation of demographic information to identify problematic respondents.

The *responsive verification protocol* consisted of multiple interactive interview steps. Step 1 – For those who indicated that they ingested household cleaners on any of the three questions, a follow-up question was presented: e.g. “You indicated you drank or gargled diluted bleach solution. Did you really drink or gargle diluted bleach solution, or did you indicate you did so by mistake on the last survey question?” Step 2 - For those who verified their response in Step 1, a follow-up question was presented to verify that this was done intentionally: e.g. “You indicated you drank or gargled diluted bleach solution. Did you engage in this cleaning behavior intentionally?” Finally, in step 3 we asked all respondents who reported ingesting household cleaners to provide more context about their answer in an open-ended format, “Please describe the steps you took to clean this way. What cleaning product did you use? How did you administer it? This research is very important for public health policy and we very much appreciate your time and input!”

For those respondents who did not report ingesting household cleaners but did report engaging in other dangerous cleaning practices steps 1-3 were followed for those practices. *Demographic verification*

Demographic information was examined for extreme outliers and implausible entries. Entries were considered extreme outliers when they were more than ten standard deviations from the population mean.

## Results

Overall, eight percent of the full sample (55/688) reported having ingested at least one of the three cleaners. 68.8% (473/688) of respondents passed the screener, and 31.2% (215/688) did not pass the screener. In line with results from Study 1, we again found that of those respondents who reported ingesting cleansers, the majority (43 of 55 or 78%) were problematic respondents. Conversely, most respondents who did not report ingesting cleanser were not problematic respondents (461/633 or 72.8%). When problematic participants were taken out of the analytic sample, reports of drinking/gargling household cleaner, soapy water and diluted bleach were 1.05%, 1.48% and 0.63% respectively.

We next examined how many respondents verified having ingested cleanser (either drinking/gargling household cleaner, soapy water, or diluted bleach) after being presented with a verification prompt. We observed that only 3 out of 12 participants did so, constituting 0.63% of those who passed the screener. For the three respondents who verified having ingested cleanser, when asked whether they engaged in these practices intentionally or unintentionally, 2 out of the 3 respondents said they did so unintentionally, leaving only one person, or 0.21% of those who passed the screener, having verified ingesting cleaner and having done so intentionally.

We next examined the pattern of demographic information provided by this respondent. Several elements of their demographic data stood out as suspicious. First, this participant reported being 20 years old and having four children. Having four children is certainly not uncommon but having four children at 20 years old is unusual, especially in the US. This respondent also reported a weight of 1900 pounds, and a height of “100”. Although they did not indicate whether they responded in centimeters or inches, either (8.3 feet or 3.2 feet) is an extreme outlier. On the open-ended question, when asked to provide more detail about having ingested cleanser, their response was “YXgyvuguhih”. Figure 3 shows the flow chart for these results.

**FIGURE 3.**
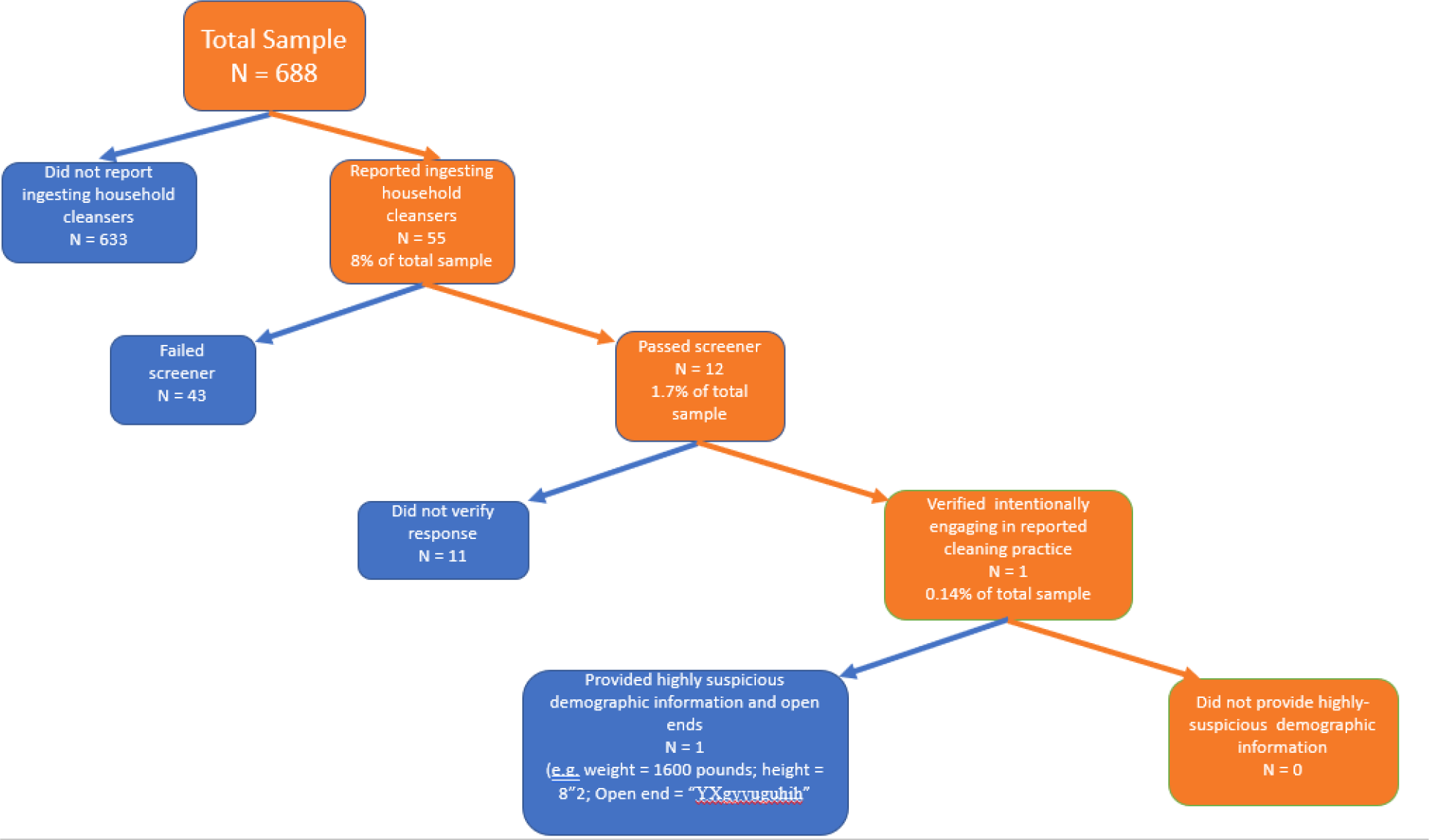
Flow chart of the step-wise process of identifying problematic responses among those who report ingesting household cleaners.

Across the entire sample, the open-ended responses further indicated that at least some respondents misunderstood the questions. For example, one respondent who reported drinking and gargling soapy water indicated on the open-ended response that, “My mother made me wash my mouth out with soap water because I was cursing, and I accidentally swallowed some”. It is clear from this response that the respondent did not read the question carefully and did not consider that; a) the question specifically inquired about practices in the time of Covid-19, and b) specifically inquired about cleanliness practices that were motivated by reducing the likelihood of Covid-19 infection, rather than something that happened to them in childhood. This specific respondent was also flagged by the attentiveness instrument. Their open-ended response provides corroborating support for flagging this respondent as problematic by the attentiveness instrument, and further demonstrates how answers to Yes/No questions cannot always be taken at face value.

Overall, we did not find a single respondent who provided any reasonable or compelling open-ended description of cleanser ingestion. This is in contrast with other cleaning practices reported on the survey. For example, regarding ‘misting the body with cleaning spray or alcohol spray after being in public spaces’, the open-ended responses were clear and detailed, e.g. “I use 70% alcohol in a spray mist bottle when I take my clothes off and I mist them, let them air dry and then put them in the washing machine.”

In general, the open-ended responses indicate that many of the questions were at times misinterpreted even by non-problematic participants. This raises the possibility that the estimates obtained for the non-ingestion practices are also inflated among non-problematic respondents. Specific examples of these open-ended responses are presented in the discussion.

With regard to the association between dangerous cleaning practices and health outcomes, we found that reported dangerous cleaning practices accounted for 14.7% of the variance in the number of reported health symptoms among respondents who did not pass the screener or did not verify ingesting cleaners, F (1, 213) = 36.7, p. < 001. However, among non-problematic respondents, less than .001% of the variance in the number of reported health symptoms was explained by reported dangerous cleaning practices, F (1, 398) = 0.556, p. = .82.

## Discussion

Society is becoming increasingly dependent on survey research to describe attitudes and behavior in multiple areas of human life including politics, medicine, social science, public health, marketing, and business. With the advent of online recruitment, it has become faster and more affordable to collect survey data than ever before. However, respondents who are non-attentive, respond randomly to survey questions, and misrepresent their true attitudes - referred to here as ‘problematic respondents’ - pose fundamental threats to the validity of data collected on surveys. Previous studies have shown that problematic respondents claim to use fictitious drugs, make claims on surveys that are contradicted by direct in-person observations, and misrepresent who they are by providing inconsistent and implausible demographic information. Such claims can make it appear that certain attitudes and behaviors (referred to here as ‘events’) are much more common than they actually are. This poses a particularly grave concern for the detection of rare events, since problematic respondents can create the illusion that almost any event can be observed in the population with a frequency of a few percentage points.

The goal of the present study was to examine whether claims that respondents make on surveys about ingesting household cleansers to protect themselves against Covid-19 infection are in large part an artifact of inattention, response error, and mischievousness - i.e. problematic respondent bias. Specifically, our main hypothesis was that reports of ingesting household cleaners are examples of illusory events that are observed on surveys due to problematic respondent bias.

Across two studies, with close to 1400 total respondents, we replicated previous findings (see Gharpure et al, 2020) showing that around 4% of respondents report engaging in each of the three cleanser ingestion activities queried in the survey - drinking or gargling household disinfectant, soap, and bleach. Eight percent of the sample reported engaging in at least one of these practices. However, consistent with the notion that problematic respondents can create the illusion that almost anything occurs in the population no matter how implausible, we also observed that 3% of respondents reported that they have never used the Internet (an answer they provided while using the internet), 5.8% reported having “suffered a *fatal* heart attack”, and 7% reported “Eating concrete for its high iron content”. These findings are consistent with a recent comprehensive report by the Pew Research Center that 7% of respondents from over 50 different opt-in panels provide “bogus” data (Kennedy et al., 2020).

After having categorized respondents into problematic and non-problematic groups based on inattention and implausible claims, we observed that 80 to 90% of reports of ingesting household cleansers were made by problematic respondents. We additionally found that misreading questions, misinterpreting the intent of questions, and respondent error accounted for the rest of such claims. Once inattentive, mischievous, and careless responses are taken out of the analytic sample, we find no evidence that people intentionally ingest household cleansers for protection against Covid-19 infection. These results strongly suggest that reports of ingesting household cleansers to confer protection against COVID-19 infection are an artifact of problematic respondent bias.

### Types of problematic respondent bias and their effects

We observed that problematic respondent bias introduces two sources of error; 1) It increases noise and, 2) it introduces systematic bias toward affirmative responses. Some problematic respondents who do not read questions tend to randomly select from among the available response options. This decreases the signal-to-noise ratio, tending to drive estimates toward the mean of the distribution. Random responding not only makes less common practices appear more common than they actually are, but also makes more common practices appear less common than they actually are. Specifically, the majority of non-problematic respondents reported having increased general non-dangerous cleaning practices to prevent Covid-19 infection. However, problematic respondents were less likely to report engaging in general non-dangerous cleaning practices than non-problematic respondents. This finding is consistent with noise attenuating estimates toward the middle of the distribution.

We also observed that bias is proportionally greater among the lowest frequency events. Specifically, we observed that problematic respondents were three times more likely to report using cleanser and disinfectant on hands or bare skin compared to non-problematic respondents. However, problematic respondents were twenty-nine times more likely to report gargling or drinking bleach solution compared to non-problematic respondents. Across all cleaning practices examined in this study, the more common practices were proportionally less likely to be affected by problematic respondent bias.

We also observed that some problematic respondents systematically select a ‘Yes’ response from among the available response options. While random responding introduces noise that is uncorrelated across items in the dataset, systematic yea-saying introduces error that is correlated across unrelated items. Evidence of systematic yea-saying among problematic respondents can be seen by examining the correlation between cleanliness practices and health outcomes. Among problematic respondents, over 25% of variance in health outcomes is explained by dangerous cleaning practices. However, among non-problematic respondents this relationship was not significant. This shows that problematic respondents systematically answer *yes* to a variety of questions across the survey, artificially driving up associations between unrelated events. These results are consistent with previous studies showing that associations between variables are dramatically reduced or eliminated altogether when problematic respondents are removed from a sample (e.g. Robinson-Cimpian, 2014).

### Which dangerous cleanliness practices are people actually engaging in?

The spread of COVID-19 across the US and the world created fear of contagion, leading people to seek ways of protecting themselves against infection. Our data is consistent with previous findings in showing that most people began to engage in more stringent cleanliness practices in order to reduce the likelihood of infection as a result. The current study explored which specific cleanliness practices people started to engage in during the COVID-19 pandemic, querying respondents about seven practices that are considered dangerous and are not recommended by the CDC.

Overall, we find that the reported rates of all seven cleanliness practices examined in this study are dramatically lower than previously thought because most reports of these practices are provided by problematic respondents. The three practices that involve ingesting household cleansers, in particular, are entirely an artifact of problematic respondent bias, misreading questions, and respondent error. At the same time, more than 5% of non-problematic respondents reported engaging in several dangerous cleaning practices, including washing food products with bleach, using cleaner or disinfectant on hands and bare skin, and misting the body with cleansers. What can be concluded about the rates at which people engage in these practices, and what are the implications of these findings for public health?

Overall, the interpretation of reports of non-ingestion related dangerous cleaning practices is complicated by open-ended responses that make it clear that some non-problematic respondents did not fully understand the intent of these questions. As is evident from open-ended responses, even otherwise attentive and well-intentioned survey-takers can misunderstand the intention of questions or may not be perfectly aware of the meaning of specific terms and phrases. For example, the meaning of the term “household cleaner” was not clear to all respondents, and at least some put regular soap in that category. To the question “Did you use household cleaner to clean or disinfect bare hands or skin?”, examples of open-ended responses included: “I washed my hands with antibacterial soap”. The use of antibacterial soap is not considered a dangerous cleaning practice. Thus, the interpretation of reports of using household cleaner on bare skin hinges on whether people categorized various non-dangerous cleaners such as soap as a “household cleaner”.

For inhaling vapors of household cleaners, there was also evidence of misinterpretation. Vapors from a wide variety of household products, including cleaners, can be inhaled. The question in this study was specifically intended to detect whether people are inhaling cleaners for the purpose of preventing infection. To achieve this goal, vapors of household cleaner would presumably be inhaled by breathing directly from an open-air container, but the current question does not specify the specific method of delivery. This led some respondents to interpret the question to mean any inhalation of cleaner, even if that cleaner was inhaled during a normal cleaning routine. For example, in response to the question “Did you inhale the vapors of household cleaners like bleach?” examples of open-ended responses included: “I poured product on the floor and began to mop”. While inhaling vapors of cleaning products during daily cleaning activities such as mopping can have negative long-term health consequences, such cleaning practices are not considered to pose an acute danger to health.

Other questions also showed evidence of misinterpretation. To the question “Did you mist the body with cleaning spray or alcohol spray after being in public spaces?”, examples of open-ended responses included: “I used 70% alcohol in a spray mist bottle when I take my clothes off and I mist them, let them air dry and then put them in the washing machine”. This respondent did mist with alcohol, but did not do so directly on the body, and then washed off the alcohol in the washing machine before their clothes made direct contact with the body. To the question of “Why did you wash fruits, vegetables, or other food products with bleach?” examples of misinterpretation included “I used soap and a special sponge to make sure I get a deep clean on the food I serve to my son”. As many open-ended responses show, people often generalize to cleaning products other than the specific one that is mentioned in the question.

Overall, for all questions, open-ended responses make it evident that the practices that at least some people were reporting on are not considered dangerous. These responses show that at least some people did not realize the importance of bleach as a specific cleaner of interest of several questions and were reporting on using cleaners like soap on their hands and skin or to clean fruits and vegetables. On other questions, respondents were affirming that they engaged in a certain practice, like misting alcohol, but did not realize the importance of other parts of the question, which focused on direct contact with the skin. Additionally, respondents did not always make a distinction between practices that they were engaged in prior to the start of the pandemic and those that they started doing specifically to prevent infection.

Given the uncertainties in the way that these questions were understood by respondents, the implications of these reported practices for public health remain unclear. It is not clear from these findings whether substantial numbers of people engage in specific dangerous practices. To fully understand the implication of these reported practices for public health, future studies should examine these practices in more detail, focusing on several details not addressed here or in previous studies: 1) effort should be made to define the specific activities and substances in the question so as to make it very clear to respondents which activities the researchers are asking about, and 2) efforts should be made to specifically define terms such as “household cleaner” so as to leave no room for doubt that the practices in question pose a health risk. Because the current study and previous studies that used these questions were not designed to provide a systematic examination of any of these practices, it remains difficult to ascertain whether the practices reported on in this survey were being practiced at all and whether they pose a substantive risk to public health.

### Public health implications

Several practices examined in this study, including the ingestion and inhalation of household cleaner have been documented in the medical literature. Such practices are most commonly observed among vulnerable populations, such as low socioeconomic groups, teenagers, and prison inmates. There have been several documented cases of blindness and death because of drinking hand sanitizer since the start of the Covid-19 pandemic. However, in virtually all cases, people ingest and inhale sanitizer for its alcohol content and psychotropic effects.

Health behavior theories emphasize that social norms can impact individual decision making and may be even more salient among vulnerable populations. Presenting practices such as the ingestion and inhalation of household cleansers as being practiced by tens of millions of people risks normalizing such practices and potentially inadvertently reinforcing them by messaging that such practices are normative. For this reason, presenting the results of surveys that are subject to problematic respondent bias is itself a matter of public health concern. The reporting of any rare event detected on a survey should be subjected to rigorous examination and should require an additional level of stringency when screening respondents.

### Comparisons with previous studies

We interpret our data to mean that previously reported results of dangerous cleaning practices to prevent Covid-19 infection (Gharpure at all, 2020), are due to problematic respondent bias. However, it is worth addressing the possibility that the differences between our finding and previously reported results are perhaps due to a slight difference in the timing of data collection or sample composition.

Our key finding was that the observed rate of practices such as ingesting bleach was exactly the same as those reported previously. Thus, the differences between our data and the previous reports are not in the reported rates of cleaning practices but in the underlying explanation of those reports.

### Recommendation for best-practices for eliminating problematic respondent bias from survey data

It has been well documented in the literature examining best practices for online research that detecting rare events requires an additional level of stringency when screening respondents. What is generally sufficient for most studies may prove inadequate when looking at low frequency events. Generally speaking, when survey data suggest that people are engaging in surprising and extremely unusual behaviors—especially those with important public health implications—it is critical to examine whether such results may have been influenced by problematic respondents prior to drawing strong conclusions from the data. Here, we recommend three practices that researchers should follow when collecting data online. These practices will improve data quality on surveys seeking to measure rare events as well as improve the signal to noise ratio in any survey.

#### Do not rely on third-party solutions without testing them first

Standard procedures used in the opt-in panel industry to protect surveys against bad data do not confer sufficient protection for the vast majority of scientific surveys. This is especially true when the goal is to detect relatively rare events. No solution is perfect, and even if a solution works to protect certain types of surveys there is no guarantee that it will work in all cases. For example, some data quality solutions employed by opt-in panels may protect against duplicate responses, bots, straight lining, and virtual private networks (VPNs) which conceal a respondent’s country of origin, but may not be effective against inattentiveness, mischievous respondents, or acquiescence bias.

#### Use response validation

It is important to follow up with individual respondents to gather more detailed information about reported behaviors, especially in the case of rare events. Researchers can ask respondents to describe these practices in an open-ended format, such as to provide specific examples of their behavior, to provide more context, and to describe the rationale for their practices. Researchers could even set up video interviews with select respondents to verify that the respondents are indeed real, that they fully understand what is being asked of them, and that their behaviors are being reported accurately. Even a handful of interviews can provide important evidence that such practices are really occurring in the population.

#### Use validated instruments

Validated screeners and other forms of intra and extra-survey data quality measures should be incorporated into all surveys as protection against problematic respondents. Throughout this paper we have described a variety of screening mechanisms that have been developed and validated by multiple research teams. Other more in-depth discussions of how to develop and incorporate data protection measures have been described elsewhere. At a minimum, researchers should choose only those tools that have been previously validated to accomplish the following goals: (a) Maximize the detection of inattention and minimize screening participants out of a survey based on other constructs related to memory, education, or culture-specific knowledge (b) Have clear, correct answers that have been previously validated to be selected by the vast majority of attentive respondents, and (c) Should not be overly stringent or too easy. The later requirement typically requires extensive testing.

#### Data quality instruments should be well-matched to the project

Researchers conducting surveys should choose data protection measures that are well-suited for the demands placed on the attention of respondents in their specific study. When choosing a data protection instrument for a specific survey, researchers should adopt a ‘fit-for purpose’ approach (see Litman, Robinson and Rosenzweig, 2020). Each data protection instrument should fit the purpose of the survey. There is no single strategy that works for all types of surveys. For example, surveys that aim to detect rare events will need higher levels of security compared to other surveys.

## Conclusions

The results of this study showed that the vast majority of dangerous health-related cleanliness practices were almost exclusively reported by problematic respondents, indicating that reports of high-risk cleaning practices to provide protection against Covid-19 infection were an artifact of problematic respondent bias. Not screening out problematic respondents causes rare events to be severely overestimated and normal practices to be underestimated. Over the last several decades our society has become increasingly dependent on survey research, with more than 80% of surveys using online respondents for at least some of the data collection (Kennedy et al., 2020). Problematic survey respondents pose a fundamental challenge to all survey research and threaten the validity of public-health policy. For this reason, it is critical to develop validated instruments to protect surveys against such bias. To mitigate against these threats surveys should rigorously check for problematic respondents using a fit-for-purpose approach, particularly when the survey aims to measure rare events. Using these techniques significantly increases the accuracy of measurement and prevents problematic respondents from invalidating survey results.

## Data Availability

Data are available upon request.

